# Automated cervix biometry, volumetry and normative models for 3D motion-corrected T2-weighted 0.55-3T fetal MRI during 2nd and 3rd trimesters

**DOI:** 10.1101/2025.04.16.25325947

**Authors:** Alena U. Uus, Agnieszka Glazewska-Hallin, Simi Bansal, Megan Hall, Charline Bradshaw, Jordina Aviles Verdera, Mary A. Rutherford, Jana Hutter, Lisa Story

**Affiliations:** Biomedical Computing Department, School of Biomedical Engineering and Imaging Sciences, King’s College London, London, UK; Research Department of Early Life Imaging,School of Biomedical Engineering and Imaging Sciences, King’s College London, London, UK; Department of Women and Children’s Health, King’s College London, London, UK; Smart Imaging Lab, Radiological Institute, University Hospital Erlangen, Germany; Fetal Medicine Unit, Guy’s and St Thomas’ NHS Foundation Trust, London, UK

## Abstract

Fetal MRI provides superior tissue contrast and true 3D spatial information however there is only a limited number of number of MRI studies investigating cervix during pregnancy. Furthermore, there are no clearly formalised protocols or automated methods for MRI cervical measurements. This work introduces the first deep learning pipeline for automated multi-layer segmentation and biometry for 3D T2w images of the pregnant cervix. Evaluation on 20 datasets from 0.55T and 3T acquisitions showed good performance in comparison to manual measurements. This solution could potentially minimise the need for manual editing, significantly reduce analysis time and address inter- and intra-observer bias. Next, we used the pipeline to process 270 normal term cases from 16 to 40 weeks gestational age (GA) range. The inlet diameter and length showed the strongest correlation with GA which is in agreement with the gradual remodeling and softening of the cervix prior to birth. We also generated 3D population-averaged atlases of the cervix that are publicly available online.

## Introduction

In clinical practice, 2D transvaginal ultrasound (TVUS) measurement of the cervical length remains the gold standard diagnostic and surveillance test in assessing the probability of preterm birth^1^. The appearance of the cervix during pregnancy has been well documented using TVUS. Ultrasound-guided cervical length (CL) measurement allows surveillance and timely therapeutic interventions such as initiation of progesterone therapy and / or cervical cerclage^1^. Normal cervical anatomy in pregnancy has a cylindrical or bell-shaped structure on ultrasound, with a length between 30 and 40 mm^2^. A CL less than 25mm in singleton pregnancies is associated with an increased risk of preterm birth^3^. Recent studies have also focused on the use of TVUS for assessment of caesarean section scars, including niches, following fully dilated caesarean section^4^ and injury following cone biopsy as part of the diagnosis/treatment of cervical dysplasia^5^.

The CL is measured by TVUS in the sagittal plane from the inlet to outlet of the cervix (internal to external cervical os)^6^. However, the measurement of CL on TVUS may be affected by multiple factors including: intra- and inter-observer variability^7^, bladder fullness, pressure exerted by the ultrasound probe, lower uterine segment contractions as well as nonlinear cervical shape and different measuring protocols between clinical centers^8^,^9^. Furthermore, although 3D ultrasound can provide volumetric data, clinical TVUS is generally limited to 2D measurements.

Magnetic resonance imaging (MRI) is non-invasive. It provides superior tissue contrast to TVUS and true 3D spatial information, allowing better differentiation between internal cervical zones^10^, including detection of cervical canal cysts. MRI-measured CL has been shown to correlate well with TVUS-measured CL^11^. Positive correlation has been noted between MR stromal signal intensity, mid-canal cross section and delivery gestation^10^,^12^ potentially related to cervical softening precluding cervical dilatation. MRI also enables the development of 3D models of the uterus, cervix and fetal membranes to study the interaction between cervical shape, position and symmetry and membrane adhesion and stiffness, culminating in a mechanical stress state of the cervix^13^. However, MRI cervical studies to date have comprised of heterogeneous acquisition and measurement protocols and relatively small sample sizes.

### Contributions

In this work, we present the first multi-label parcellation protocol for 3D motion-corrected T2w SSTSE MRI images of the cervix imaged during pregnancy. This protocol is used as a baseline for training a deep learning pipeline for automated cervix segmentation of 3D DSVR-reconstructed cervix images based on semi-supervised training. The segmented cervix layers are used to automatically generate a standard cervix length biometry measurement. The feasibility of the wider application of the pipeline is then tested based on segmentation of 270 normal 0.55T and 3T T2w fetal MRI datasets for generation of volumetry and biometry gestational charts of the cervix across 16-40 weeks GA range. We also present the first population-averaged 3D fetal MRI atlases of the normal cervix anatomy that are publicly available online.

## Methods

### Cohorts, datasets and preprocessing

The fetal MRI cohort used in this work includes 330 datasets from 16 to 40 weeks gestational age (GA) acquired at St. Thomas’ Hospital, London. The datasets were collected prospectively as part of the “PiP” (REC 16/LO/1573), “PRESTO” (REC 21/SS/0082), “NANO” (REC 22/YH/0210), “MiBirth” (REC 23/LO/0685) and “MEERKAT” (REC 21/LO/0742) studies. All experiments were performed in accordance with relevant guidelines and regulations. Informed written consent was obtained from all participants.

The T2w single-shot turbo spin echo (ssTSE) stacks in the datasets were acquired with 2 different acquisition protocols:

- 156 datasets acquired on 0.55T Siemens MAGNETOM Free.Max MRI system with with 6-element flexible coil and a 9-element spine coil built into the patient table using a dedicated low field fetal acquisition protocol^14^ with TE=105ms, acquisition resolution 1.48 x 1.48mm, slice thickness 4.5mm, 0mm gap and 3-6 whole uterus stacks;
- 174 datasets acquired on 3T Philips Achieva MRI system using a 32-channel cardiac coil with TE=180ms, acquisition resolution 1.25 x 1.25mm, slice thickness 2.5, 0mm gap and 2 whole uterus stacks;

The inclusion criteria were: singleton pregnancy; no overt (e.g., niches) structural anomalies in the maternal cervix anatomy; absence of severe acquisition artifacts; sufficient number of stacks with full cervix coverage; clear visibility of the cervix in reconstructed images; supine position of the mother during scanning. The entire cohort (used for training, testing and normative ranges) consisted of 330 datasets including both normal control and cases with high preterm birth risks, pre-eclampsia, and various fetal anomalies. The datasets were obtained from particpants with maternal age (21-47 years) and body mass index (BMI) (17-40). The normal control cervix cohort with 270 datasets included only pregnancies with term delivery without reported fetal or maternal complications and with a cervical length greater than 2.0cm (based on the ultrasound measure)^15^).

The 3D images of the cervix were reconstructed using the in-house pipeline based on classical DSVR method^16^ in SVRTK toolbox with a uterus sagittal stack as a template (since this plane is conventionally used for cervical measurements) and automated deep learning masking. The output images have 0.8mm isotropic resolution.

### Definition of cervix parcellation and biometry protocol

The protocol for cervix segmentation and biometry was defined by clinicians (LS, AGH, SB) with extensive experience in fetal and cervical MRI. The parcellation labels were manually defined using ITK-SNAP^17^ in 3D DSVR images of the cervix. The segmented regions include 3 main cervical layers: endocervical canal, inner stroma, outer stroma (similarly to^12^,^18^) and any cysts if present. The selection criteria for these labels were based both on the relevance to clinical quantitative volumetry studies as well as clear visibility in 3D T2w DSVR cervix images. The length and diameter cervix biometry measurements were defined based on conventional ultrasound guidelines^19^. The final protocol illustrated in Fig. 1 is summarised as follows:

**Figure 1.**
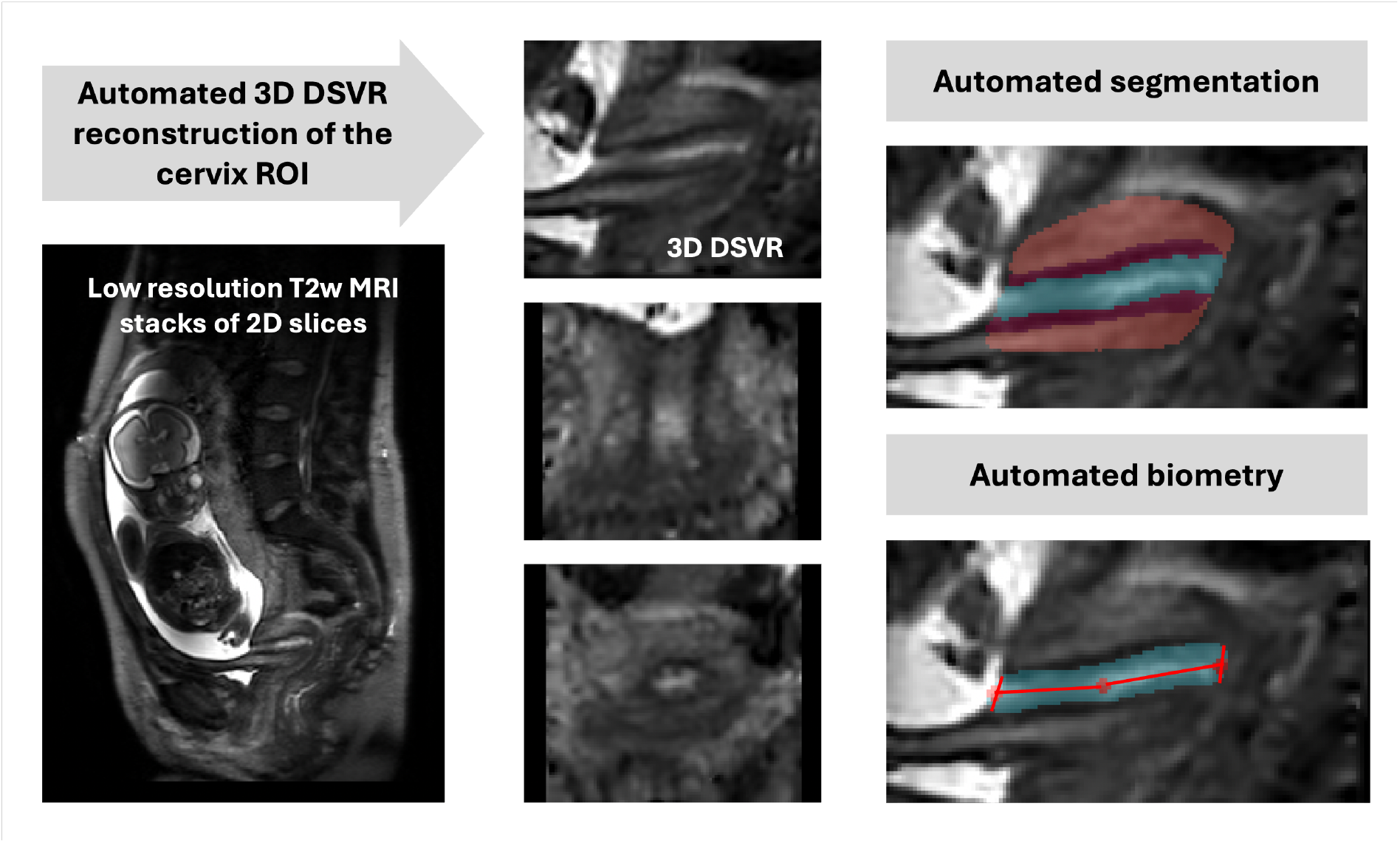
The proposed pipeline for 3D T2w MRI fetal cervix analysis: automated DSVR reconstruction, 3D segmentation (volumetry) and biometry.

- The endocervical canal (ECC): the internal cervical mucosa ROI characterised by a mixture of regional high and low signal intensity corresponding to varying properties of the mucus / mucus plug;
- The inner stroma or subglandular zone (SGZ): the internal layer of the cervical tissue that encloses the canal and is characterised by low T2w signal intensity;
- The outer stroma (OS): the external layer of the cervix that encloses the inner stroma and is characterised by intermediate signal intensity slightly lower than the surrounding soft tissue;
- The fluid-filled cysts: focal regions of hyperintensity predominantly located within the canal;
- The inlet diameter of the cervix: diameter of the cervical canal at the interface between the uterus and cervical opening measured in the mid-sagittal plane with respect to the inlet;
- The outlet diameter of the cervix: diameter of the cervical canal at the level of the anterior and posterior lip of the cervix measured in the mid-sagittal plane of the outlet;
- The cervical length: the sum of two lines drawn between the centrepoints of the inlet and the outlet and the midpoint of the cervical canal.

### Population-averaged 3D T2w atlases of the cervix

In order to formalise the average normal cervix model during pregnancy, we created 2 population-averaged 3D T2w cervix atlases from two acquisition protocols from 81 (0.55T, TE=105ms) and 61 (3T, TE=180ms) 3D DSVR reconstructed images of normal control subjects without reported anomalies. The atlases were generated in MIRTK toolbox in 5 iterations using rigid, affine and non-rigid registration followed by averaging with Laplacian sharpening with 0.8mm isotropic resolution. The atlases were then segmented by the proposed 3D segmentation network described below. The atlases and the proposed parcellation protocol are publicly available online at the KCL fetal MRI cervix atlas repository (https://gin.g-node.org/kcl_cdb/cervix_mri_atlas).

### Automated segmentation and biometry of the cervix

The proposed solution for automated 3D parcellation of the cervix in 3D DSVR MRI images is based on the classical multi-label 3D UNet segmentation^20^ in MONAI^21^. The training was performed in three stages using semi-supervised approach. At first, we used a set of 20 manually segmented 3T and 0.55T DSVR cervix images (5 labels: EEC, SGZ and OS layers, cysts, bladder) to pretrain the preliminary version of the network. The manual segmentations were done by AGH and SB in ITK-SNAP based on the defined protocol. The training was performed for 10000 iterations with MONAI-based augmentation and additional MIRTK histogram matching. The pretrained network was then used to automatically segment the next set of 30 images with variations in cervical anatomy (e.g., diameters, length, canal intensity, etc.) that were manually refined by AGH, SB and AU and used in the next of stage of training. Next, the pipeline was employed to segment the final set of 50 0.55T and 3T DSVR images that were manually refined by AGH, SB and AU. The final version of the network was trained using 100 training and 10 validation datasets for 100000 iterations.

As illustrated in Fig. 1, the automated biometry step for measurement of the cervical length and canal diameters is based on extraction of inlet, outlet and midpoint landmarks from the segmented cervix labels. At first, the inlet and outlet ROIs are localised as interfaces between the canal (including cysts) and background. Next, the centre-point of the inlet and outlet are computed as averages of the corresponding ROIs. The cervix midpoint is computed as the average coordinate of the canal label. This is followed by fitting of two lines through the inlet, midpoint and outlet to compute the cervical length in mid-sagittal plane as a sum of two lines similarly to clinical practice^19^. The inlet and outlet canal diameters are measured in the sagittal plane as fitted lines along the corresponding inlet and outlet centre-point levels with the boundaries defined by the inner layer. The inlet and outlet diameters were therefore measured in different sagittal planes located according to their corresponding centrepoints.

### Implementation and evaluation details

The cervix segmentation code based on MONAI and python scripts for biometry along with the trained models are publicly available in 3D Fetal MRI GitHub repository (https://github.com/SVRTK/auto-proc-svrtk). The full segmentation pipeline and the automated 3D reconstruction can be executed using the publicly available standalone CPU-based docker at the SVRTK Docker repository (https://hub.docker.com/r/fetalsvrtk/svrtk; tag: *general_auto_amd*).

The parameters of the employed 3D UNet are as follows: five encoder-decoder blocks (output channels 32, 64, 128, 256 and 512), convolution and upsampling kernel size of 3, ReLU activation, dropout ratio of 0.5, batch normalisation, and a batch size of 1. We employ AdamW optimiser with a linearly decaying learning rate, initialised at 1×10-3, default beta parameters and weight decay 1×10-5. The training was performed using combined Dice and Cross Entropy Loss The preprocessing steps included resampling with padding to a 128x128x128 grid. We used standard MONAI augmentation including bias field, contrast adjustment and affine rotations (+/- 45 degrees).

The proposed cervix analysis pipeline was tested on 20 datasets from 21-40 weeks gestation with 0.55T and 3T acquisition protocols, varying image quality and varying anatomy. Automated segmentations were evaluated by comparison of the original network outputs vs. manual refinement the DL labels in terms of ICC, Dice in addition to the absolute and relative volume difference. The refinement of the DL labels was performed by AU in ITK-SNAP. Automated biometry was evaluated by comparison of automated outputs vs. average of manual measurements performed by by four observers (AGH, SB, CB, AU) in terms of ICC, absolute and relative differences in mm. We also computed inter-observer ICC for manual measurements.

### Generation of normal cervix volumetry and biometry charts

We used the proposed deep learning pipeline to segment 270 3D DSVR cervix images from normal control fetal MRI datasets from 16-40 weeks GA range and 0.55T and 3T acquisition protocols. All segmentations were reviewed and manually refined, if required. They were then used to run automated biometry for all cases. Based on the extracted measurements, we performed an Analysis of Covariance (ANCOVA) to examine the relationship between fetal GA, maternal parameters (age, height, ethnicity, weight, BMI) and cervix measurements, while adjusting for confounding effects.

## Results

### Population-averaged 3D T2w atlases of the cervix

The created average 3D cervix MRI atlases at 0.55T and 3T field strength are shown in Fig. 2. They have continuous and visible layer structure. The atlases were inspected by LS, AGH and SB and the anatomy was confirmed to be correct and corresponding to the normal cervix during pregnancy. The cervix layer segmentations have clear boundaries at tissue interfaces. Notably, despite the different tissue contrast and different subjects, the atlases at 0.55T and 3T have similar appearance in terms of the tissue interface boundaries, relative positioning, shape and proportions between the labels.

**Figure 2.**
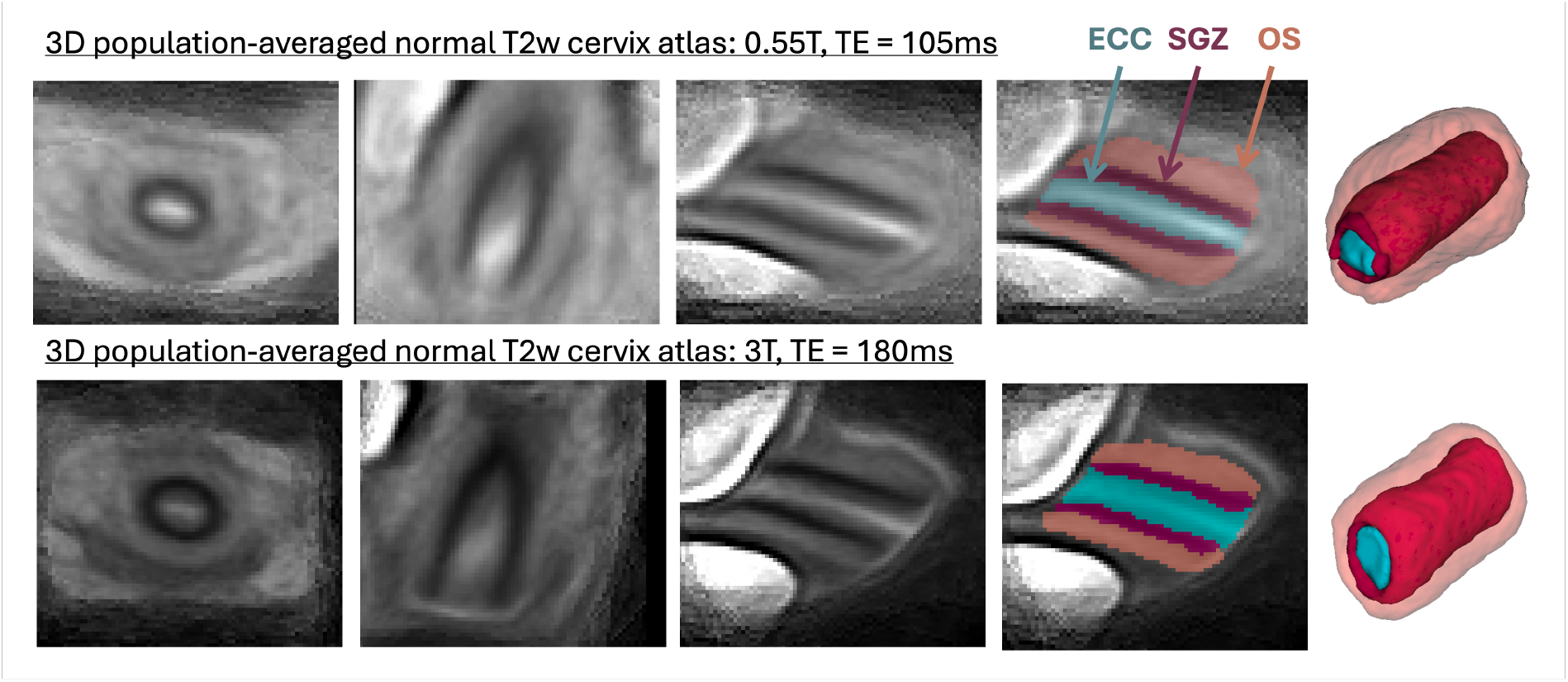
3D population-averaged T2w atlases of the cervix at different field strengths and the corresponding segmentations of the cervix layers.

### Evaluation of automated segmentation and biometry of the cervix

The results of the assessment of the automated segmentation and biometry performance on 20 test cases from 21-40 weeks GA range and 2 acquisition protocols are summarised in Fig. 3. The UNet was able to detect all ROIs including cysts in all test subjects. All cervix layers have smooth boundaries corresponding to the changes in signal intensities and cervix anatomy. While comparison of the automated label volumes with manually refined labels showed high intraclass correlation coefficient (ICC) it also showed *<* 10% differences in the canal and inner layer ROIs. This was caused by slight systematic overestimation of the inner layer by the network due to high variance in intensity differences between cases. Notably, manual refinement took less than 3 minutes per case vs. 20-40 minutes normally required for manual segmentation.

**Figure 3.**
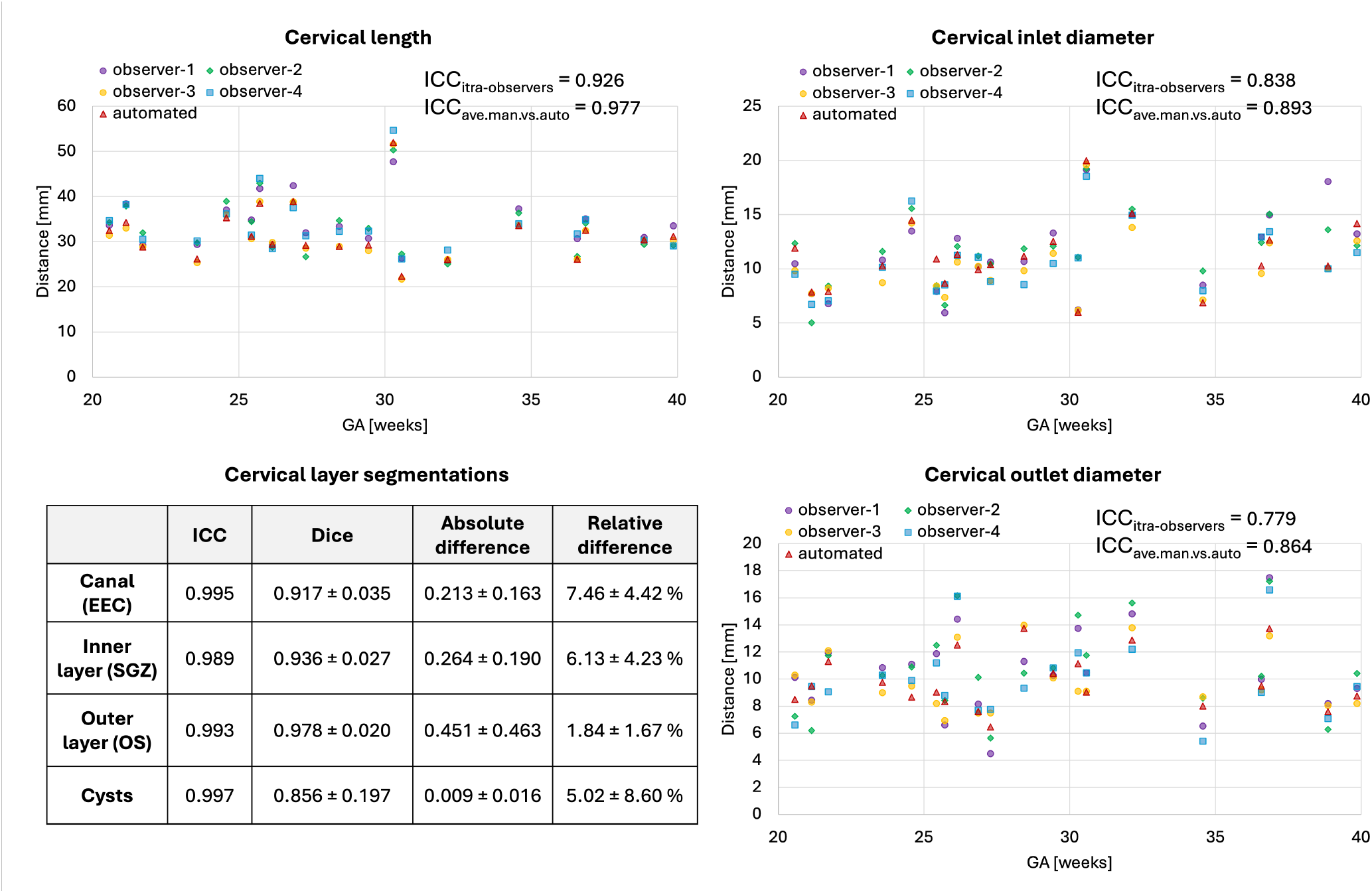
Results of testing of the proposed automated cervix analysis pipeline on 20 cases: segmentation of the layers (A) and biometry (B) vs. manual labels and measurements.

The plots of biometry measurements in Fig. 3 show a high degree of variance in the manual biometry measurements between four observers with the inter-observer ICC lower than ICC between averaged manual vs. automated biometry results. This highlights the known inter-observer variability in 2D measurements^7^. One of the factors contributing to this variability was the challenge in defining the location of the inlet and outlet centrepoints and differentiation between the canal and the interface of the inner layer. The automated biometry landmarks were inspected for all 20 test cases and confirmed to be acceptable. The average absolute errors between the average manual vs. automated measurements were within 1.56 *±* 1.06, 1.18 *±* 0.81 and 0.99 *±* 0.84 ranges for the length, inlet and outlet diameters, correspondingly.

While this evaluation confirms the general feasibility of automation of cervical volumetry and biometry it also demonstrates the need for further optimisation of the pipeline particularly for parcellation of the canal and inner layer and development of automated quality control.

### Normal control cervix volumetry and biometry charts during pregnancy

Next, we used the proposed pipeline to process 270 normal control term 0.55T and 3T MRI datasets from 16-40 weeks GA (Fig.5). The volumetry and biometry charts generated from the corresponding automated cervix segmentations are presented in Fig. 6. Only minor segmentation refinements were required in 37% cases (predominantly in the canal label due to varying signal intensities and poorly defined outlet and inlet interfaces) which did not produce significant difference in the global volumetry trends. There was a notable variance in both volumes and biometry measurements for the whole GA range indicating the normal variance in the anatomy within the healthy control pregnancies . Similarly to inter-patient differences in length and diameters, the shape of the cervix in the normal control cases also showed a wide range of variations. E.g., Fig. 4 demonstrates the segmentation examples for cases with varying anatomy and from different field strengths. In addition to varying length and diameter, there were variations in composition of the canal content, thickness and intensity of the inner layer, utero-cervical angle and curved shape/bending of the cervix. In 36% of all cases, cysts were detected and predominantly located within the inlet region. Cysts were significantly less visible in 0.55T datasets. For the global volumetry trends the cysts volume was included into the total canal volume. Segmentation of the outer layer was generally characterised by higher uncertainty due to lack of visibility of the interface boundary.

**Figure 4.**
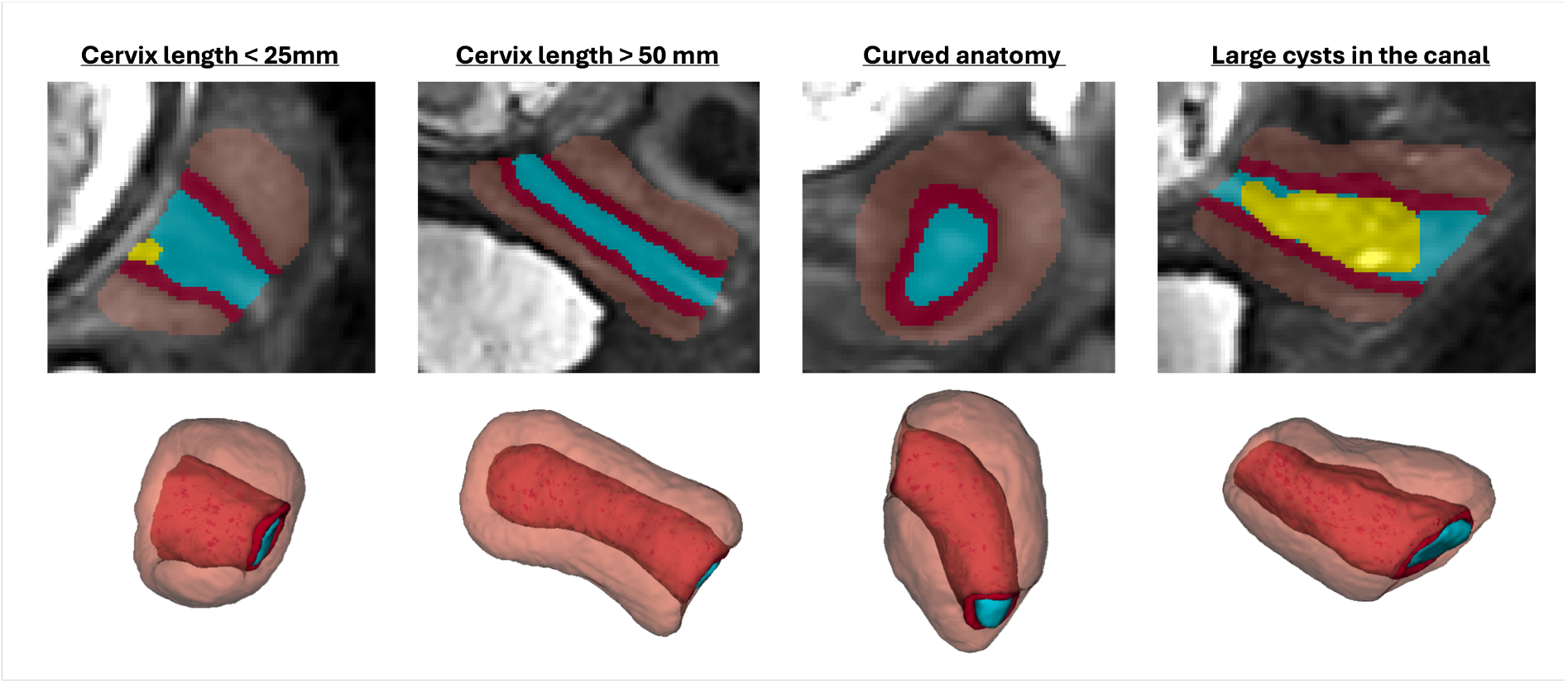
Examples of predicted cervix segmentations for 3D T2w fetal MRI reconstructions of cervix cases with anatomical variations.

**Figure 5.**
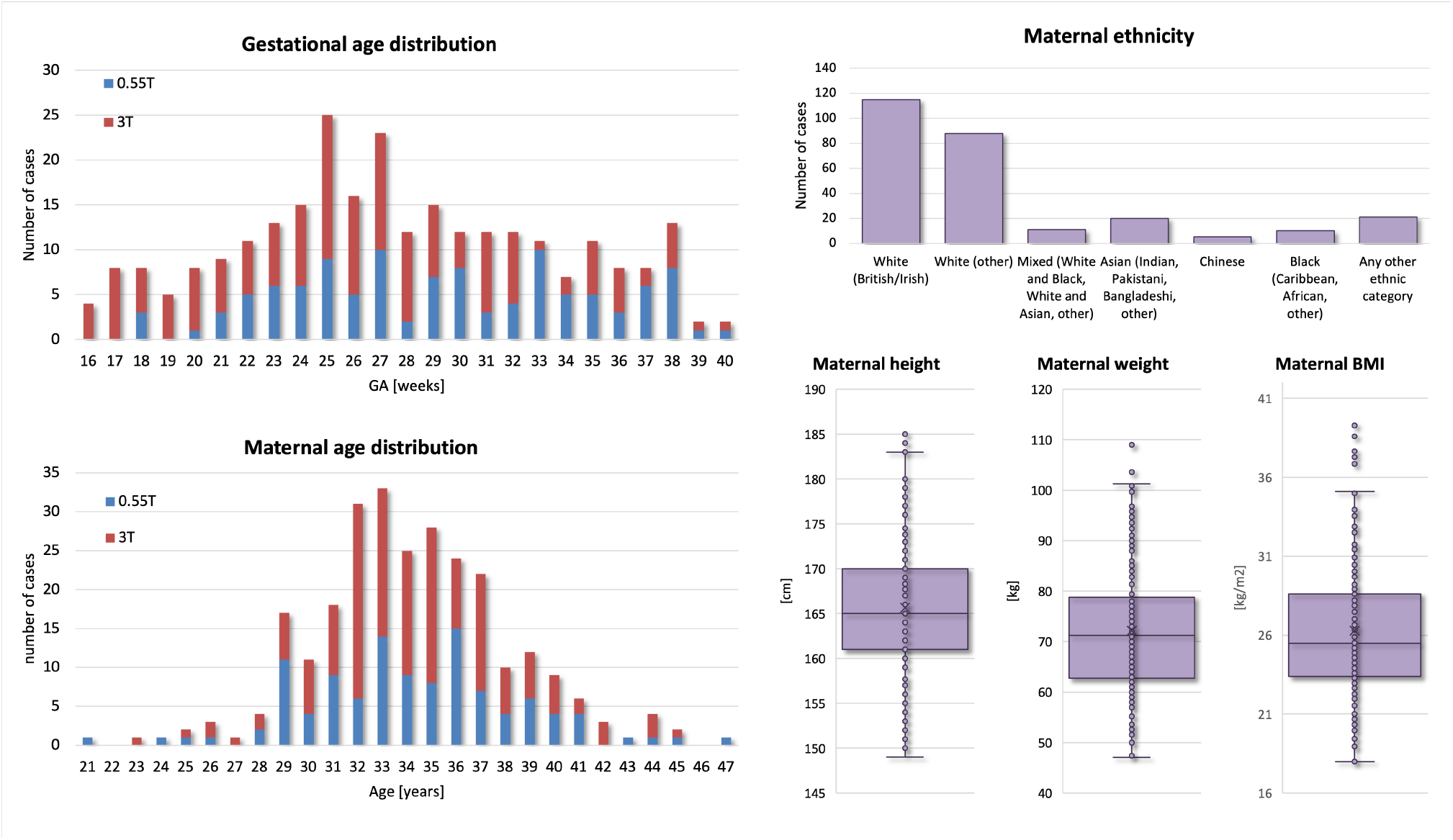
Summary information of 270 normal control cohort datasets used for analysis of cervical changes during 2nd and 3rd trimesters.

**Figure 6.**
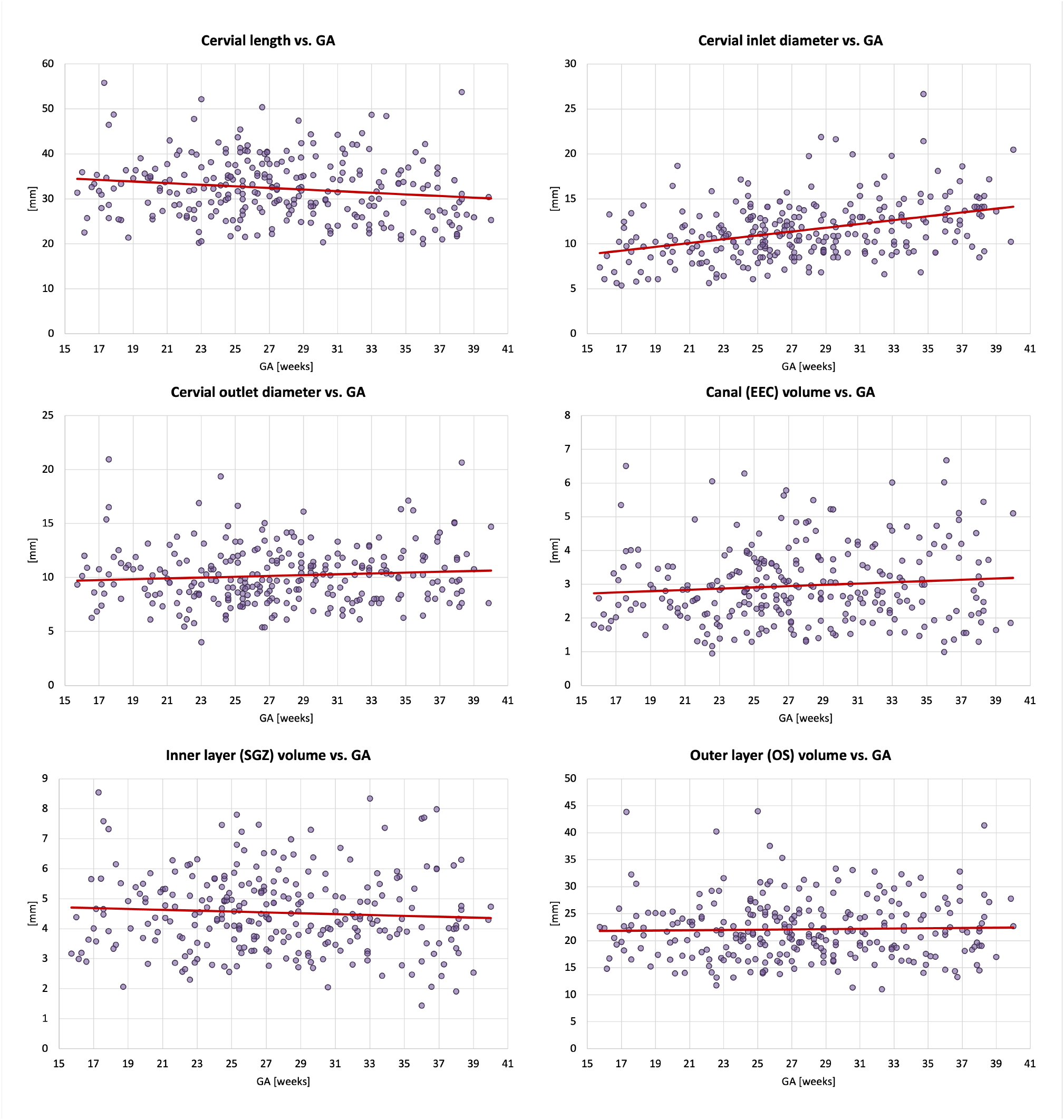
Biometry and volumetry growth charts generated from 270 normal control 3D reconstructed 0.55T and 3T T2w fetal MRI datasets.

The global trendlines in the graphs and ANCOVA analysis show significant correlation in cervical length (*p <* 0.01) and inlet diameter (*p <* 0.0001) with GA. The decrease in values can be attributed to gradual cervical remodeling (shortening and effacement) and softening of the cervical layer tissue as the body prepares for labor as well as increasing mechanical pressure from the weight of the fetus. In addition, the inner layer volume (*p <* 0.01) and cervix outlet diameter (*p <* 0.05) were also significantly affected by GA with slightly decreasing trends. Notably, while the inclusion criteria did not include cases with the length below 20 mm the decreasing cervical length trend could have been more pronounced at later gestation with the addition of larger numbers of control cases. This suggests that the normal cervical length range at later gestation could be lower than during the 2nd trimester. Fig. 7 show examples of 0.55T and 3T longitudinal datasets with pronounced changes in the cervix anatomy shape between three timepoints. The decreasing cervix length and increasing inlet and outlet diameters with GA follow the global trends in the charts. Yet, the changes in volumes are not consistent between the cases which could be related to inter-patient variations as well as limitations of segmentation quality and tissue visibility.

**Figure 7.**
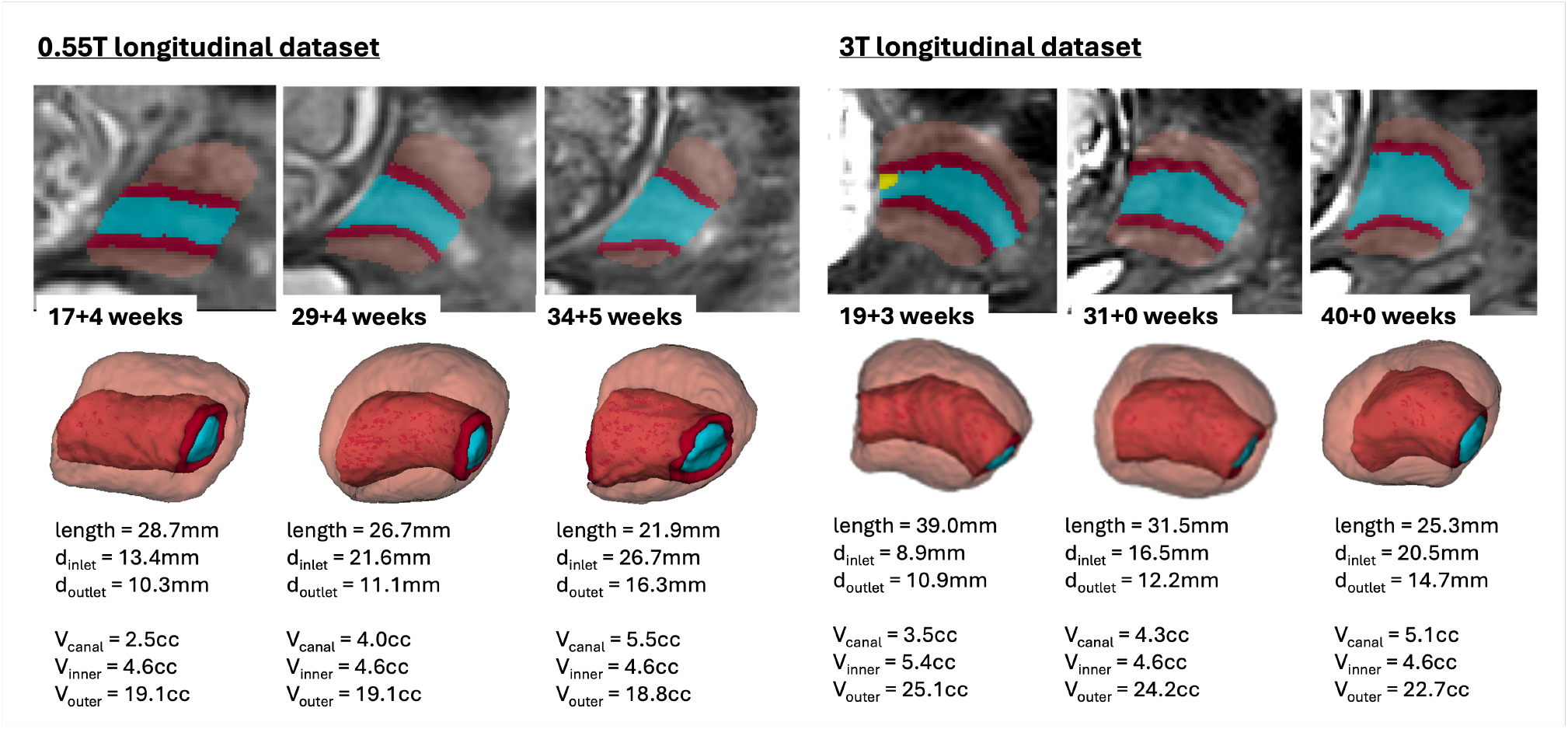
Examples of changes in the cervix anatomy in 0.55T and 3T longitudinal datasets with 3 time points: segmentation labels and 3D models and automated measurements.

In terms of the impact of maternal parameters on cervical measurements, maternal age showed significant correlations with the inner and outer layer volumes (*p <* 0.05), cervical length (*p <* 0.05) and cervix inlet diameter (*p <* 0.01). In the current cohort, older age is associated with higher volumes, longer cervix and smaller inlet diameter. Yet, maternal age was predominantly higher than 28 years which limits this analysis. After correction for GA and other factors, there was no correlation between cervix measurements and maternal height and weight. The diversity in the predominantly white ethnicity cohort was not sufficient for comprehensive evaluation.

There was a significant impact of the scanner field strength on the canal volume and outlet diameter (*p <* 0.001) values that can be attributed to pronounced differences in tissue layer contrast (e.g., Fig. 2).

## Discussion

In this work, we introduced the first formalisation of multi-label cervix parcellation protocol for 3D T2w MRI during pregnancy together with automated solutions for cervix segmentation and biometry. We also presented 3D population-averaged atlases of the normal cervix anatomy at 3T and 0.55T MRI field strength that are publicly available online together with the parcellation protocols.

Definition of the cervix layer labels relevant to quantitative volumetry studies was performed based on both T2w signal intensities and visibility of anatomical interfaces in 3D DSVR-reconstructed images by clinicians with extensive experience in cervix and fetal MRI. Measurements of the cervix length and diameters for the biometry protocol follow the conventional clinical practice for MRI and ultrasound. Next, based on the proposed protocols and manual labels we trained a 3D UNet network for segmentation of the cervix into three layers with an additional label for cysts on 100 datasets from 0.55T and 3T datasets. The cervix biometry step is based on extraction on inlet, inner centrepoint and outlet landmarks from the canal label followed by automated linear measurements. Quantitative evaluation of the automated segmentation network and biometry pipeline based on comparison with manual labels and measurements demonstrated fairly robust performance for different field strengths. The main challenges for the proposed pipeline were segmentation of the interface between the canal and inner layer due to varying regional mucus intensity ranges. Segmentation of the outer stroma layer was also characterised by high degree of uncertainty due to the limited contrast and visibility with the external tissue interface. This also indicates that automated segmentation and biometry outputs should always be reviewed and fine-edited, if required. Comparison between the automated and average manual biometry measurements from four observers showed relatively good agreement. The lowest ICC was for the outlet diameter due to challenges in tissue interface localisation. Notably, the inter-observer biometry ICC was lower than ICC between average manual vs. automated. This is in agreement with the generally known inter-observer variability in cervical measurements and stresses the need for automated quality and certainty control of measurements.

Next, we used the networks to run automated volumetry and biometry measurements of 270 normal term control cases (*<*20mm length) during second and third trimesters from 0.55T and 3T acquisitions. Only minimal manual refinement was required in less than 37% of cases. This is a significant improvement in terms of time (<3 minutes per case) vs. the conventional manual segmentation approach (20-40 minutes per case) as well as 3D continuity of layer ROIs.

Analysis of the cervical parameters at different gestational ages demonstrated high variance in the normal anatomy for all variables. The inlet (internal os) diameter had the highest correlation, increasing with GA. The trend of cervical length decreasing with GA may be underestimated, given that we did not include cases under 20mm length, which may be a non-pathological finding for some women. These findings are consistent with the expected gradual shortening and softening of the cervix with gestation. This correlation was also present in examples of longitudinal dataset with 3 timepoints. The association we have presented of older maternal age and a longer cervix is consistent with the literature^22^. The finding of higher volume and shorter inlet in older women is new. However, the current cohort did not have sufficient representation of mothers younger than 29 years old. The cohort ethnicity was also not sufficiently diverse for detailed analysis, which emphasizes the need for a targeted study.

These proof-of-concept results confirm the general suitability of deep learning for automated analysis of the cervix and this is the first step towards automated quantitative assessment of cervical anatomy during pregnancy for large scale MRI studies.

## Limitations and future work

While this paper showed promising results for normal cervical anatomy, the pipeline has not been trained or tested on abnormal cases. Our future work will focus on optimisation of segmentation and biometry protocols and network for abnormal anatomy including funneling, scars, short cervix, as well processing of not-pregnant cervix scans and further refinement of segmentations at tissue interfaces. There is also a potential benefit in training classification networks for automated quality control as well as automated image-based detection of pathological and high-risk cases.

We are also planning to use the proposed methodology to compare normal and high-risk cohorts (e.g. preterm birth) to detect anomalies antenatally. We will further optimise the segmentation protocol and extend the list of biometrics with the angular, crossectional canal area and stroma area measurements^12^. Another potential direction is application of the automated segmentations for computation of mean diffusion parameters^18^) and 3D simulation-based structural modelling^13^.

## Conclusions

In this work, we introduced the first automated pipeline for multi-layer segmentation and biometry of motion corrected T2w 3D MRI images of the cervix. It was used to generate normal volumetry and biometry cervix charts during 16-40 weeks GA range and for different acquisition protocols. The results demonstrated robust performance of the pipeline for different gestational ages and acquisition protocols with only minor manual refinements required. This suggests the potential feasibility of using automated measurements for large-scale quantitative volumetry studies that would significantly reduce the need for time-consuming manual input and minimise inter- and intra-observer variability. We also generated the first population-averaged atlases of the cervix at 0.55T and 3T that are publicly available online. We have demonstrated MR changes in the cervix based on gestational age. Our future work will focus on optimisation of the protocol for abnormal cases and automated image-based classification for detection anomalies and analysis of high-risk cases.

## Acknowledgements

We thank everyone who was involved in acquisition and analysis of the datasets at the Department of Perinatal Imaging and Health at Kings College London and St Thomas’ Hospital. We thank all participants and their families.

This work was supported by NIHR Advanced Fellowship awarded to Lisa Story [NIHR30166], by the Wellcome Trust, Sir Henry Wellcome Fellowship to Jana Hutter [201374/Z/16/Z], DFG Heisenberg funding [502024488], by the UKRI, FLF to Jana Hutter [MR/T018119/1], the MRC grant [MR/X010007/1], the MRC grant [MR/W019469/1], the Wellcome Trust and EPSRC IEH award [102431] for the iFIND project [WT 220160/Z/20/Z], the NIH Human Placenta Project grant [1U01HD087202-01], the Wellcome/ EPSRC Centre for Medical Engineering at King’s College London [WT 203148/Z/16/Z], the NIHR Clinical Research Facility (CRF) at Guy’s and St Thomas’ and by the National Institute for Health Research Biomedical Research Centre based at Guy’s and St Thomas’ NHS Foundation Trust and King’s College London.

The views expressed are those of the authors and not necessarily those of the NHS, the NIHR or the Department of Health.

## Author contributions

A.U. and A.G.H. contributed equally to this work and the manuscript. A.U. implemented the automated segmentation and biometry pipeline, processed the datasets, created the atlas and produced the graphs. A.G.H. formalised the parcellation protocol, prepared training datasets and analysed the results. S.B. prepared training datasets and analysed the results. M.H., C.B. participated in acquisition and analysis of the datasets. J.A.V. participated in acquisition of the datasets. M.R. and J.H. provided fetal MRI datasets, participated in analysis of the results and supervised various parts of the project. L.S. provided datasets, formalised the segmentation protocol, participated in analysis of the results and supervised the research. All authors reviewed the manuscript.

## Data availability

The individual fetal MRI datasets used for this study are not publicly available due to ethics regulations. The created population-averaged 3D T2w atlases of the cervix are publicly available online at KCL CDB MRI atlas repository: https://gin.g-node.org/kcl_cdb/cervix_mri_atlas.

